# COVID-19 vaccine effectiveness against SARS-CoV-2 infection in the United States prior to the Delta and Omicron-associated surges: a retrospective cohort study of repeat blood donors

**DOI:** 10.1101/2022.04.15.22273412

**Authors:** Eduard Grebe, Elaine A. Yu, Marjorie D. Bravo, Alex Welte, Roberta L. Bruhn, Mars Stone, Valerie Green, Phillip C. Williamson, Leora R. Feldstein, Jefferson M. Jones, Michael P. Busch, Brian Custer

**Author notes:** Senior authors (Michael P. Busch, Brian Custer).

## Abstract

To inform public health policy, it is critical to monitor COVID-19 vaccine effectiveness (VE), including against acquiring infection. We estimated VE using a retrospective cohort study among repeat blood donors who donated during the first half of 2021, demonstrating a viable approach for monitoring of VE via serological surveillance. Using Poisson regression, we estimated overall VE was 88.8% (95% CI: 86.2–91.1), adjusted for demographic covariates and variable baseline risk. Time since first reporting vaccination, age, race-ethnicity, region, and calendar time were statistically significant predictors of incident infection. Studies of VE during periods of Delta and Omicron spread are underway.

## INTRODUCTION

Coronavirus disease 2019 (COVID-19) vaccines have a critical role in preventing symptomatic illness, including serious disease, caused by severe acute respiratory syndrome coronavirus 2 (SARS-CoV-2) [1], and in reducing viral transmission [2]. COVID-19 vaccines approved or authorized for emergency use in the US by the Food and Drug Administration had high efficacy against severe disease in Phase III trials, ranging from 67% to 95% [1, 3-5]. However, long-term management of the SARS-CoV-2 pandemic remains a substantial challenge, especially as variants of concern emerge. To inform public health policy, it is critical to monitor real-world effectiveness of COVID-19 vaccines at the population-level over longer periods of time [6]. Numerous individual-level and environmental factors (e.g., local community transmission, social distancing and other mitigation practices, vaccine type, and vaccination coverage) impact vaccine effectiveness (VE), underscoring the importance of and need for population-based studies of VE against SARS-CoV-2 infection.

Evaluating VE is challenging given the multitude of endpoints relevant to SARS-CoV-2 infection. Many COVID-19 VE studies have evaluated severe endpoints, such as mortality and hospitalizations, often in higher risk populations. Mild and asymptomatic infections likely account for the vast majority of SARS-CoV-2 infections, with serosurveys indicating that many more infections occur than diagnosed cases [7]. Vaccine breakthrough infections are frequently asymptomatic with one study finding that among >10,000 breakthrough infections, over a quarter were asymptomatic [8]. Asymptomatic individuals (including with vaccine breakthrough infections) can transmit SARS-CoV-2 to others, and thus preventing asymptomatic infection is important to decrease widespread community transmission [9]. Therefore, VE against acquiring infection is important to assess.

Constraints on large-scale population-level monitoring of VE include the cost and logistical challenges of enrolling and following cohorts of vaccinated and unvaccinated individuals, and laboratory assays. Molecular diagnostic assays, such as reverse transcription polymerase chain reaction (rtPCR) assays applied to respiratory swab samples, are the gold standard for diagnosing SARS-CoV-2 infection. However, molecular diagnostic assays have a limited detection window, necessitating frequent follow-up that reduces the feasibility of ongoing large-scale surveillance, higher cost, and lower throughput. In contrast, serological assays that detect binding antibodies (Abs) against the Spike (S) and Nucleocapsid (NC) viral proteins are lower cost, higher throughput, and can identify SARS-CoV-2 infections after resolution.

We estimated VE using a retrospective cohort study among repeat blood donors who donated during the first half of 2021 at Vitalant, a major US blood collection organization (BCO). This proof-of-concept study demonstrates a viable approach for continued near real-time monitoring of VE via serological surveillance among repeat blood donors.

## METHODS

Beginning in June 2020, Vitalant tested all donors for anti-S and anti-NC Abs using the Ortho VITROS anti-SARS-CoV-2 S Total Ig and Roche Elecsys® NC Anti-SARS-CoV-2 assays. For this analysis, we included donations from donors who donated at least twice between January 1 to July 6, 2021, with anti-S and anti-NC test results. Donors self-reported any COVID-19 vaccination (vaccine type and number of doses not specified) at each donation via the Donor Health Questionnaire (DHQ) and each interdonation interval was categorized as vaccinated or unvaccinated time at risk, based on serological and COVID-19 vaccination status. Specifically, we defined an interdonation interval as vaccinated time at risk if the donor was vaccinated and had anti-S Abs but not anti-NC Abs at the first timepoint and unvaccinated time at risk if the donor was not vaccinated at both timepoints and had no anti-S Abs at the first timepoint. Interdonation intervals where donors were infected (anti-NC positive) at the start were excluded. Additionally, intervals during which vaccination took place were excluded because the proportion of vaccinated versus unvaccinated time could not be determined. A single donor could contribute both unvaccinated and vaccinated time at risk. Exact dates of vaccination and vaccine manufacturer were not recorded on the DHQ responses, although preliminary data from a survey of Vitalant blood donors conducted subsequently indicates that more than 90% received an mRNA vaccine (Moderna or Pfizer-BioNTech). Incident SARS-CoV-2 infections were identified using anti-NC seroconversion; vaccine breakthrough infection defined as seroconversion after self-report of COVID-19 vaccination.

We used multivariable regression to evaluate the association between incident SARS-CoV-2 infection rate and COVID-19 vaccination and reported adjusted IRR (aIRR). The final model adjusted for the calendar month during which a donor’s follow-up ended (to control for variable baseline risk), time vaccinated at the start of the final observation interval (to control for time-dependent protection), sex, age, race-ethnicity, and the Vitalant-defined geographic region in which donations were collected. Time at risk was treated as an offset in the regression analysis, with uninfected donors contributing the full follow-up time, and infected donors contributing half of the interval immediately during which infection occurred and full intervals during which infection did not occur. A log link function was used, and analysis conducted in R version 4.1.2 (R Foundation for Statistical Computing, Vienna, Austria). VE was defined as 1-aIRR. Since a single donor could contribute both unvaccinated and vaccinated exposure time, and the resulting within-donor correlation would not be reflected in standard errors from generalized linear models without a donor random effect, we computed 95% confidence intervals (CIs) using 10,000 iterations of donor-level bootstrapping (resampling donors with replacement). As a comparator, we used naïve person-time methods to estimate incidence (number of events/time-at-risk) in vaccinated and unvaccinated donors, reported unadjusted IRRs, and calculated VE as 1-IRR.

During the study period, vaccine availability and uptake increased rapidly in the blood donor population and SARS-CoV-2 incidence declined rapidly during the early months of 2021. This raised a concern of potential bias in VE estimates caused by unvaccinated time at risk accumulating disproportionately during a period when baseline risk of acquiring the infection was higher, which could lead to an overestimate of the protective effect of vaccination. We conducted an extensive sensitivity analysis using simulated data to evaluate bias across VE estimation approaches and inform model selection. We simulated 1,020 datasets similar to the Vitalant repeat donor dataset, with varying rates of declining incidence, rates of increasing vaccination, and proportions of the population vaccinated by the end of the period. We evaluated several VE estimation methods and data selection rules, including unadjusted incidence rate ratios (IRRs) based on conventional person-time methods (naïve analysis), Cox proportional hazards regression on a calendar timescale, and Poisson regression adjusted for calendar time; methods were assessed for absolute and relative bias and precision (see Supplementary Appendix).

## RESULTS

In total, 61,618 donors who donated in 18 geographic regions across the United States contributed 407,449 unvaccinated person-weeks and 326,752 vaccinated person-weeks of time at risk during the study period (January 1 to July 6, 2021). Most donors (77%) contributed a single interval (median 1, range 1–25), and the median interval length was 56 days (IQR: 28–71). Poisson regression adjusted for the calendar time when a particular donor’s follow-up ends (either with an infection event or still at risk) had low bias, good precision, and confidence interval coverage >95% (see Supplementary Appendix).

We identified 1,653 incident infections in unvaccinated donors, and 100 vaccine breakthrough infections. Kaplan-Meier survival curves for both groups are shown in Figure 1. Using multivariable Poisson regression, we estimated overall VE was 88.8% (95% CI: 86.2–91.1), derived from an aIRR of 0.145 (95% CI: 0.116–0.180), adjusted for demographic covariates and variable baseline risk (Table 1). The number of days that the donor had been vaccinated at the start of the final interval was significant and protective. Age, race-ethnicity, geographic region and the month during which a donor’s follow-up ended were statistically significant predictors of incident infection, though sex was not (Supplementary Table 1). The naïve analysis yielded an unadjusted VE estimate of 92.5% (90.8%–93.8%, Table 1), although our simulation study found a substantial risk of bias.

**Figure 1:**
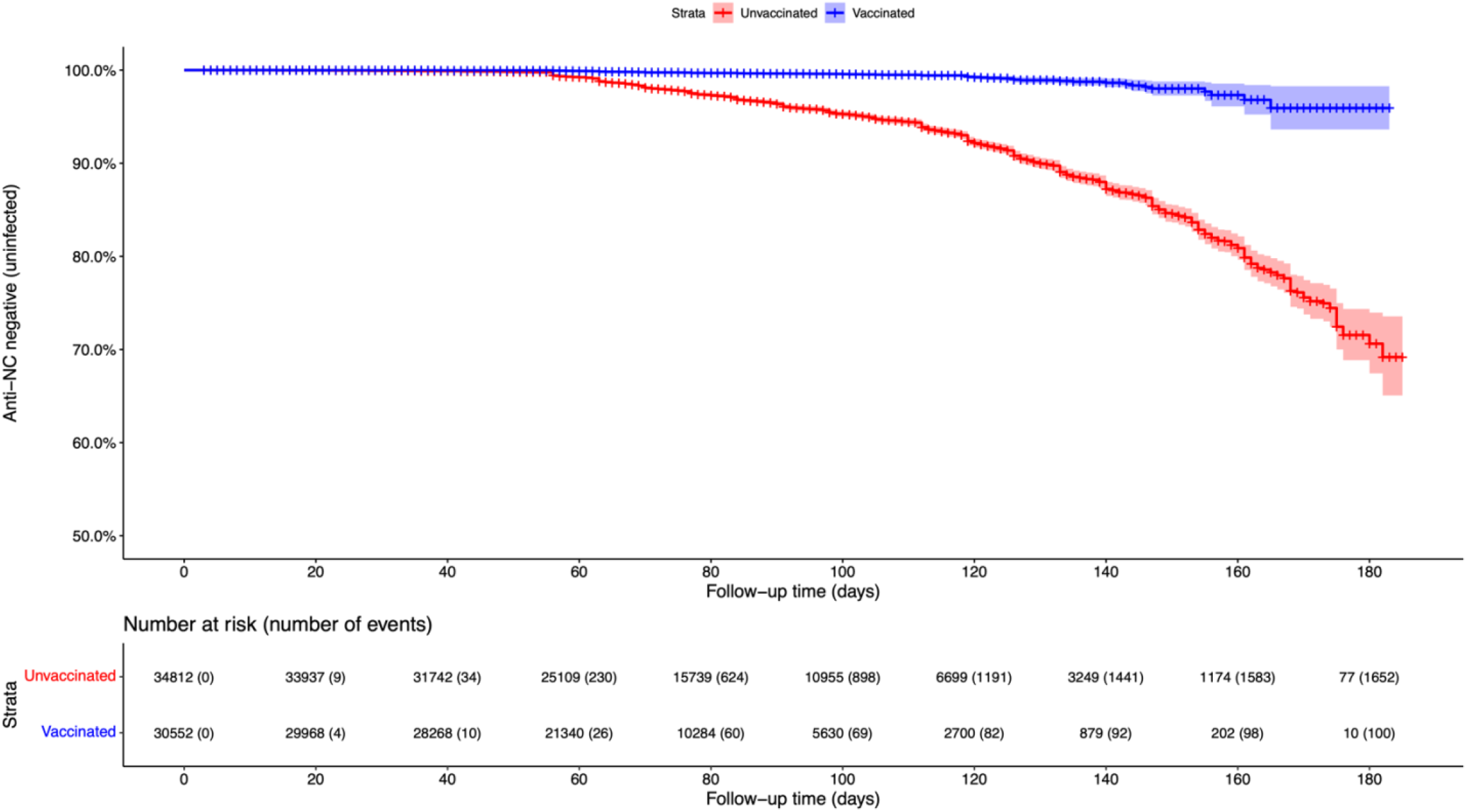
Kaplan-Meier survival curves by vaccination status comparing time to anti-nucleocapsid antibody seroconversion among US blood donors, January-July 2021.

**Table 1:** Vaccine effectiveness estimates against anti-nucleocapsid antibody seroconversion* using person-time and multivariable regression approaches among US blood donors, January-July 2021.

| **Model** | **Data inclusion period** | **Incidence in vaccinated donors**<br>***infections/10<sup>4</sup> person-weeks***<br>**(95% CI)** | **Incidence in unvaccinated donors**<br>***infections/10<sup>4</sup> person-weeks***<br>**(95% CI)** | **Incidence Rate Ratio**<br>**(95% CI)** | **Vaccine Effectiveness %**<br>**(95% CI)** |
| --- | --- | --- | --- | --- | --- |
| Poisson regression** | 1 Jan–6 Jul 2021 |  |  | 0.112 (0.089–0.138) | 88.8 (86.2–91.1) |
| Naïve person-time*** | 1 Jan–6 Jul 2021 | 3.1 (2.5–3.7) | 40.6 (38.6,42.6) | 0.075 (0.062–0.092) | 92.5 (90.8–93.8) |

\*Vaccine breakthrough infections defined as anti-nucleocapsid seroconversion after any self-report of previous COVID-19 vaccination.
Number of vaccine doses not collected. Analysis included 100 vaccine breakthrough infections.
\*\*Adjusted for age, sex, race-ethnicity, geography, and calendar time of end of follow-up (to account for variable baseline risk over time); 95% confidence intervals are obtained from donor-level bootstrapping to account for within-donor correlation in cases where a donor contributed both unvaccinated and vaccinated exposure time.
\*\*\* Unadjusted for covariates.

## DISCUSSION

VE against acquiring any serologically identifiable SARS-CoV-2 infection, was high in blood donors during the first half of 2021 at 88.8%. Our VE estimate was comparable to previous VE estimates in studies evaluating protection against SARS-CoV-2 infection based on detection of viral RNA in swab samples, which ranged from 73% to 98.2% [10-12], and mostly lower than estimates of protection against hospitalization, which ranged from 88.0% to 95.1% [10, 13, 14]. Increasing protection with longer times since vaccination in these data are consistent with maturing immune responses during the early period of vaccine implementation, but over longer timescales waning protection would be expected, as has been observed in other studies [10].

A major strength of this study was the broad assessment of a large number of repeat blood donors residing in 18 states during universal screening for SARS-CoV-2 Abs. Our model accounted for spatiotemporal factors and provided a more generalizable estimate of VE in the US population compared to previous studies in clinical populations. Our simulation-based exploration of estimation methods identified a robust statistical approach, specifically based on the criterion of having low bias in situations of complex infection and vaccination dynamics. The study constitutes an important proof of concept for the use of serological surveillance of blood donors to monitor VE over time as variants have and continue to emerge and vaccine-induced Ab responses wane or are enhanced through booster vaccinations. This approach further facilitates larger sample sizes and consequently improved precision in VE estimates at lower cost than conventional cohort studies.

Several limitations need to be considered when interpreting these results. The serological assays employed in this study have high sensitivity and specificity for past infection and vaccination-induced Abs, but lower sensitivity than molecular assays for acute infection. We did not have detailed information on COVID-19 vaccination timing, number of doses, or vaccine type, and there is the potential for misreporting of vaccination status by donors on the DHQ. To limit the impact of potential misclassification of vaccination status, we required serological evidence of a vaccination response (presence of anti-S Abs and absence of anti-NC Abs) to corroborate self-reported vaccination, although donors could still have been classified as vaccinated after receiving only the first of a two-dose series or not long enough after receiving a single-dose vaccine or the second dose of a two-dose series to have developed robust Ab responses and be considered fully vaccinated. Sporadic presentation for donation, which for some donors may be very infrequent, is inherent to repeat blood donation datasets. We developed methods to account for interval censoring of both vaccination and infection events, but some data were excluded because vaccination status or the ordering of vaccination and infection events was uncertain during the interval. We used Vitalant region as a proxy geographic variable, however this may not capture with adequate granularity the heterogeneity in local transmission. Blood donors are not representative of the US population–e.g., racial/ethnic minorities are underrepresented, and donors tend to be healthier than the general population–but there is no reason to expect that these differences are more pronounced in vaccinated or unvaccinated donors. Lastly, these data were collected before the widespread circulation of the Delta and Omicron variants of SARS-CoV-2. The Omicron variant is known to be associated with significant immune escape and reduced VE [15].

Our results showed a high VE against acquiring SARS-CoV-2 infection during the first half of 2021, although data collection preceded epidemic surges driven by spread of the Delta and Omicron variants during the second half of 2021. We have established repeat donor cohorts for continued monitoring in partnership with another large BCO and the US Centers for Disease Control and Prevention for long-term serosurveillance, including comparison of VE during periods of Delta and Omicron predominance. SARS-CoV-2 antibody assays will be complemented with donor surveys regarding COVID-19 vaccinations (primary and booster), COVID-19 diagnoses, and symptoms. This next study phase will also include estimation of VE by vaccine manufacturer and timing of primary and booster doses, as well as assessment of the severity of vaccine breakthrough infections.

## Data Availability

Blood donation data are not publicly available, but any reasonable requests to the authors will be afforded due consideration, in line with applicable blood center policies and legislation.

## Ethics and informed consent

Blood donors provided consent for the use of donation data and biospecimens in research at the time of donation. Consistent with the policies and guidance of the University of California San Francisco Institutional Review Board, Vitalant Research Institute self-certified that use of the deidentified data in this study does not meet the criteria for human subjects research. Centers for Disease Control and Prevention (CDC) investigators reviewed and relied on this determination as consistent with applicable federal law and CDC policy (45 C.F.R. part 46, 21 C.F.R. part 56; 42 U.S.C. § 241[d]; 5 U.S.C. § 552a; 44 U.S.C. § 3501).

## Disclaimer

The findings and conclusions in this report are those of the authors and do not necessarily represent the official position of the CDC.

## SUPPLEMENTARY APPENDIX

We conducted a simulation study to evaluate vaccine effectiveness (VE) estimation methods and data inclusion rules with respect to bias and precision, with a particular focus on the bias that may arise in observational studies when both incidence and the proportion of the population vaccinated changes rapidly during the study period. During the first half of 2021, the proportion of blood donors who were vaccinated increased from close to zero to approximately 80%, while over the same period incidence declined rapidly from a peak around the 2020/2021 holiday period, and then stabilized prior to the Delta variant-driven outbreak. In this situation, unvaccinated follow-up time accumulates disproportionately during a period when background incidence (and therefore baseline risk of infection acquisition) was relatively high, and vaccinated follow-up time disproportionately during a period of relatively low incidence. Methods that do not appropriately account for variable baseline risk would likely be biased and result in an overestimate of VE.

We simulated 1,020 datasets that reflected donations during a 360-day period (*t* = −180 to *t* = 180), with vaccines implemented during the second 180 days (from *t* = 0). The simulations represent an idealized scenario of rapid vaccine uptake and rapid incidence decline during the period of vaccine uptake in order to pressure-test data selection rules and estimation methods. The first 180 days are included in the simulation in order to effectively reflect the entire history of the pandemic in this simulated population without the need to specify sensible initial conditions, and mirroring the one-year period of universal SARS-CoV-2 antibody testing by Vitalant from June 2020 to early July 2021.

Each dataset consisted of 200,000 donation intervals with the time of the first donation drawn from a uniform distribution [−180,180], and the length of the donation interval drawn from a truncated normal distribution (*μ* = 65, *σ* = 35), with 90% of donations set to be from whole blood donors with a minimum inter-donation interval of 56 days, and 10% from apheresis donors with a minimum inter-donation interval of 7 days. All intervals had a maximum length of 180 days. Intervals that ended after *t* = 180 were excluded from analysis. Incidence during the first 180 days (i.e., preceding the period of interest during which vaccination was implemented) was a function of time designed to reflect multiple epidemic waves, culminating in a fixed peak incidence at day 0 of 50 infections per 10,000 person-weeks (see Figure A1). During the period of interest, incidence was an exponentially declining function of time, eventually plateauing at 20% of peak incidence (i.e., at 10 cases per 10,000 person-weeks). The function for incidence during the pre-vaccination period was a simple spline function, while the exponential decline during the period of interest is given by the following equation:

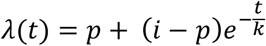

with *t* time, *p* the plateau value or asymptote, *i* the initial value, and *k* the exponential rate. The exponential rate of decline was varied stochastically in each simulation, drawn from a uniform distribution [40,80]. The proportion of the donor population that was vaccinated was kept fixed at zero until day 0, after which it exponentially increased (using the same function as above), at a rate also drawn from the uniform distribution [40,80], and plateauing at 80% (Figure A2).

**Figure A1:**
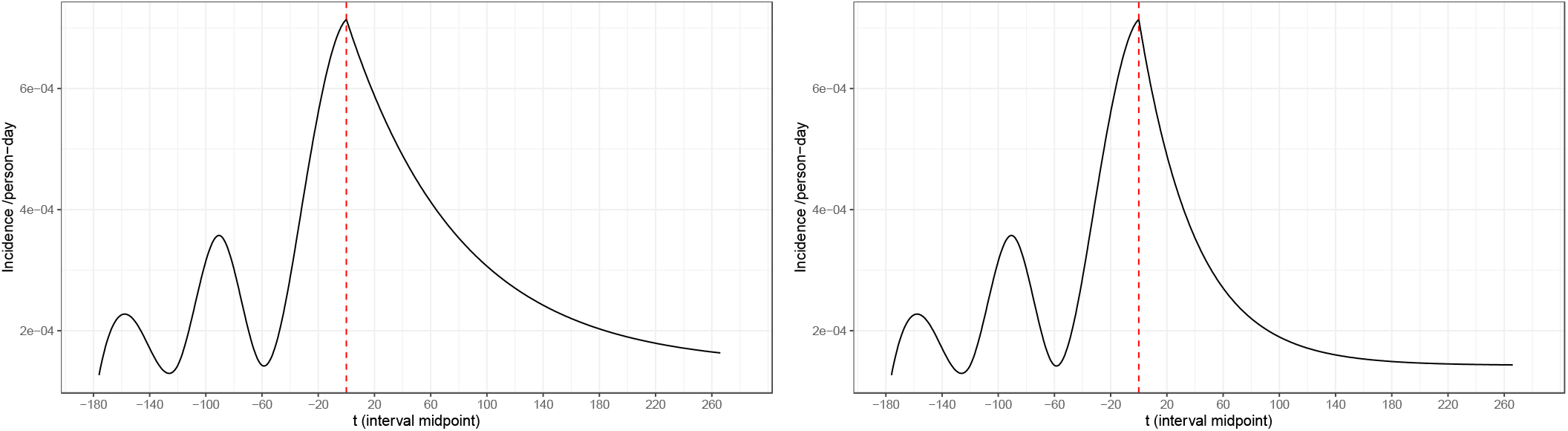
Incidence of SARS-CoV-2 infection as a function of time in simulated scenarios. *Panel A: Slower incidence decline; Panel B: Faster incidence decline*.

**Figure A2:**
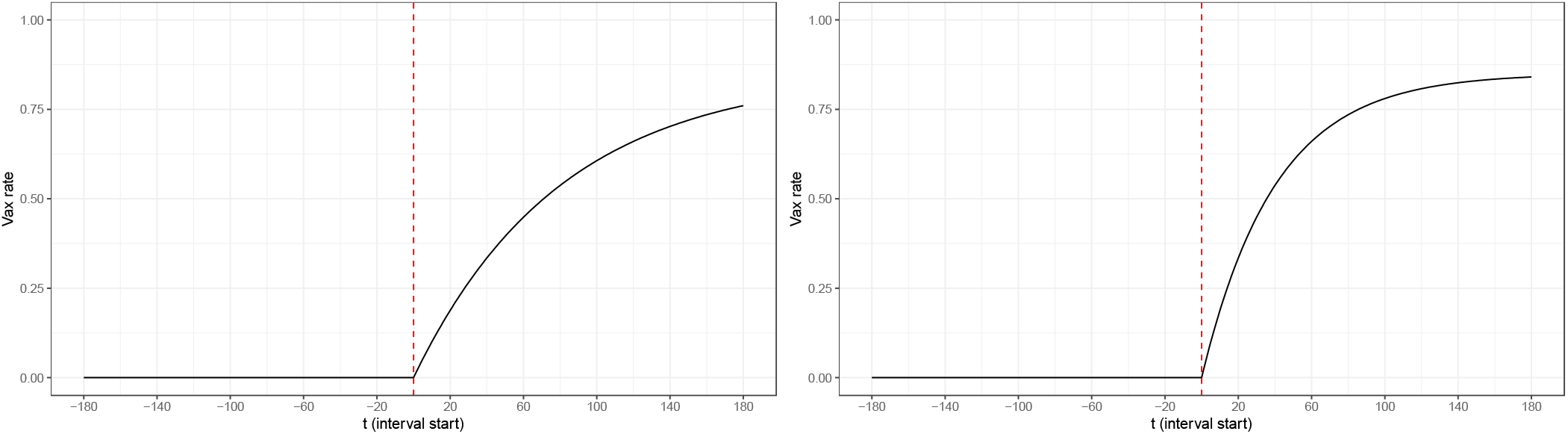
Proportion of the population vaccinated as a function of time in simulated scenarios. *Panel A: Slower vaccine uptake; Panel B: Faster vaccine uptake*.

In unvaccinated donors, the incidence function was used to compute a probability of infection occurring during an interval by evaluating the incidence function at the midpoint of the interval (expressed in infections per person-day) multiplied by the size of the interval to obtain a total risk of infection over the course of the interval. There is a loss of granularity compared to formally computing a cumulative probability of infection during an interval by integrating over time-varying incidence over the interval, but we did not believe this loss of resolution to be relevant for the purposes of this simulation study. This probability was then used to draw whether an infection took place from a binomial distribution.

Similarly, the probability that the donor was vaccinated at the start of the interval (*t*_1_) was obtained by evaluating the vaccination proportion function at *t*_1_, and the probability at the end of the interval by evaluating the function at *t*_2_, given that the donor was unvaccinated at *t*_1_, with vaccination status at *t*_1_ and *t*_2_ set binomially. If vaccination occurred during the interval, the status would be different at *t*_1_ and *t*_2_.

Vaccine efficacy was set for each simulation run at a value between 30% and 90% (drawn from a uniform distribution), and then applied to the infection probability computed as in unvaccinated donors. The probability of infection during unvaccinated and vaccinated intervals *t*_1_ to *t*_2_ are therefore given by:

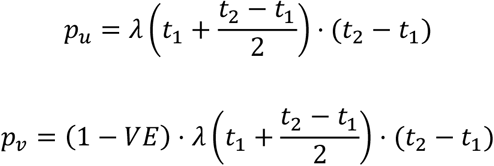

The first critical question faced in data analysis is which data points to include. In the case of these repeat donor data, we sought to determine: (1) whether to include data from the pre-vaccination period in analysis and (2) how best handle intervals during which a donor’s vaccination status changed. The latter concern arises from the fact that these data are interval censored, so that the time of an event is not precisely known – only whether it had already occurred at the time of donation. This is of particular concern since vaccination had a strong time dependence and was rapidly increasing over the period, and therefore an unobserved vaccination event during an interval would not have had a uniform likelihood to have occurred at all times in the interval. If a donor was vaccinated during an interval and did not become infected during that interval, it is not clear how to allocate time-at-risk in incidence calculations. If a donor was both vaccinated and became infected during an interval, it cannot be known in which order the events occurred. We evaluated the following strategies with respect to intervals during which vaccination took place:

- No data exclusion – consider intervals during which vaccination took place unvaccinated exposure time;
- Exclude intervals during which vaccination and infection took place, and allocate half of intervals during which vaccination but no infection took place as unvaccinated exposure time and half as vaccinated exposure time;
- Exclude all intervals during which vaccination took place;

We considered a method based on estimating incidence in vaccinated an unvaccinated donors, computing an incidence rate ratio (IRR) and then calculating VE as 1-IRR. This method does not account for variable baseline risk and cannot be adjusted for confounding (such as demographic differences associated with risk in the vaccinated and unvaccinated groups). We further considered two regression approaches: Cox proportional hazards regression, which is a time-to-event approach frequently used for studies of this kind, and Poisson regression, which is frequently used for incidence analysis when adjustment for covariates is necessary. Cox models were fit to data on a calendar timescale. Poisson models treated exposure time as an offset (the entire interval if no infection took place, and half the interval if infection took place) and used a log link. While the Cox model allows for an unspecified time-varying baseline hazard, and therefore theoretically should not be exposed to the potential bias arising from vaccinated exposure time accumulating disproportionately during a lower-incidence period, we tested both Cox and Poisson models with and without calendar time as a covariate, to control for varying baseline hazard.

In general, data selection rules that kept only intervals that started during the vaccination era (i.e. *t*_1_ ≥ 0) had lower bias. It is also intuitively apparent that additional unvaccinated exposure time during a period when no vaccinated exposure time accumulated does not add relevant information to the analysis. We therefore concluded that using only intervals with a first donation on or after 1 January 2021 was appropriate. In these simulated scenarios, the approach to handling intervals during which vaccination took place had a small effect. However, since we used incidence at the time of the midpoint of an interval to determine the probability of infection during the interval, an equal allocation of these intervals to vaccinated and unvaccinated exposure time may appear safer than it would be in reality during rapidly changing incidence. We therefore decided to exclude all intervals during which vaccination took place from analysis. It remains a potential limitation that half of an interval during which infection took place was allocated as time at risk in analysis, given that in these simulations incidence at the midpoint determined the probability of infection during the entire interval, while in a situation of changing incidence risk would be distributed non-uniformly during intervals.

We found that unadjusted IRRs derived from a conventional person-time incidence estimation approach exhibited significant bias and had poor confidence interval coverage: mean relative bias of 17%, and the confidence interval containing the true value in only 35% of the 1,020 simulated datasets. Cox regression without controlling for calendar time as a covariate performed even worse with mean relative bias of 46% and CI coverage of 0%. Poisson regression without controlling for calendar time had mean relative bias of 16% and CI coverage of 35%, pointing to the fact that Poisson regression is analytically equivalent to a crude IRR calculation when not controlling for covariates. Cox regression and Poisson regression controlling for calendar time, by including either the time of the midpoint or end of the interval as a predictor, had reduced bias and improved CI coverage. For Cox regression using the end of the interval, mean relative bias was 14% and CI coverage 62%. For Poisson regression using the end of the interval, mean relative bias was 8% and CI coverage was 84%. Poisson regression which included the midpoint of the interval performed even better (CI coverage 91%), although this improved performance is likely an artefact of the use of midpoints to determine infection risk in the simulations. In unfavorable scenarios (rapid incidence decline and slow vaccination growth, which increases the potential for bias), Poisson regression controlling for calendar time achieved CI coverage >95%, indicating that this approach effectively addresses confounding related to changing baseline hazard.

## SUPPLEMENTARY TABLES

**Supplementary Table 1:** Adjusted incidence rate ratios (IRRs) for vaccination status and covariates from multivariable Poisson regression.

| Variable | Adjusted Incidence Rate Ratio (95% CI)* |
| --- | --- |
| **Vaccination status** |  |
| Unvaccinated | *ref* |
| Vaccinated | **0.11 (0.09,0.14)** |
| **Days vaccinated at start of final interval** | **0.98 (0.97,0.99)** |
| **Sex** |  |
| Female | *ref* |
| Male | 0.99 (0.90,1.09) |
| **Age (in years at first donation)** | **0.99 (0.99,0.99)** |
| **Race-ethnicity** |  |
| White, Non-Hispanic | *ref* |
| Black, Non-Hispanic | 0.67 (0.34,1.06) |
| Asian/Pacific Islander, Non-Hispanic | **0.56 (0.31,0.84)** |
| Native American/Alaskan, Non-Hispanic | 0.70 (0.22,1.27) |
| Hispanic ethnicity | 0.93 (0.76,1.11) |
| Missing/Unknown/Refused | 1.24 (0.88,1.65) |
| **Vitalant Regional Center\*\*** |  |
| Albuquerque, NM (ALB) | *ref* |
| Billings, MT (BIL) | 1.3 (0.92,1.85) |
| Cheyenne, WY (CYS) | 1.28 (0.78,1.91) |
| Denver, CO (DEN) | **1.39 (1.10,1.80)** |
| El Paso, TX (ELP) | 1.42 (0.95,2.06) |
| Fargo, ND (FAR) | **2.00 (1.56,2.63)** |
| Lafayette, LA (LAF) | **1.55 (1.13,2.12)** |
| Las Vegas, NV (LAS) | 1.16 (0.86,1.59) |
| Lubbock, TX (LBB) | 1.25 (0.88,1.77) |
| McAllen, TX (MCA) | 0.90 (0.38,1.59) |
| Memphis, TN (MEM) | 1.38 (0.94,1.98) |
| Phoenix, AZ (PHX) | 1.08 (0.85,1.49) |
| Rapid City, SD (RAP) | 1.48 (0.98,2.04) |
| Reno, NV (RNO) | 0.82 (0.56,1.18) |
| Sacramento, CA (SAC) | **0.59 (0.44,0.79)** |
| San Francisco, CA (SFO) | **0.30 (0.17,0.47)** |
| Spokane, WA (SPK) | **1.51 (1.14,2.03)** |
| Ventura, CA (VTA) | **0.43 (0.27,0.62)** |
| **End of follow-up** |  |
| January 2021 | *ref* |
| February 2021 | 0.55 (0.32,1.20) |
| March 2021 | **0.31 (0.19,0.65)** |
| April 2021 | **0.39 (0.24,0.83)** |
| May 2021 | **0.36 (0.22,0.76)** |
| June/July 2021 | **0.26 (0.16,0.55)** |

\* Adjusted incident rate ratios in bold are statistically significant, i.e., 95% confidence interval excludes 1.
\*\* Geographic collection areas for Vitalant regional centers are shown in Figure 1 of Vassallo, RR, Bravo MD, Dumont LJ, Hazegh K, Kamel H. Seroprevalence of Antibodies to SARS-CoV-2 in US Blood Donors. medRxiv 2020.09.17.20195131; doi: 10.1101/2020.09.17.20195131.

## Notes

### Competing Interest Statement

Vitalant Research Institute receives research funds and reagents for studies from Ortho Clinical Diagnostics and Roche.

### Funding Statement

This study was funded by Vitalant.

### Author Declarations

Blood donors provided consent for the use of donation data and biospecimens in research at the time of donation. Consistent with the policies and guidance of the University of California San Francisco Institutional Review Board, Vitalant Research Institute self-certified that use of the deidentified data in this study does not meet the criteria for human subjects research. Centers for Disease Control and Prevention (CDC) investigators reviewed and relied on this determination as consistent with applicable federal law and CDC policy (45 C.F.R. part 46, 21 C.F.R. part 56; 42 U.S.C. 241[d]; 5 U.S.C. 552a; 44 U.S.C. 3501).

## REFERENCES

1. Tregoning JS, Flight KE, Higham SL, Wang Z, Pierce BF. Progress of the COVID-19 vaccine effort: viruses, vaccines and variants versus efficacy, effectiveness and escape. Nature Reviews Immunology 2021; 21:626–36.

2. Singanayagam A, Hakki S, Dunning J, et al. Community transmission and viral load kinetics of the SARS-CoV-2 delta (B.1.617.2) variant in vaccinated and unvaccinated individuals in the UK: a prospective, longitudinal, cohort study. The Lancet Infectious Diseases 2022; 22:183–95.

3. Polack FP, Thomas SJ, Kitchin N, et al. Safety and Efficacy of the BNT162b2 mRNA Covid-19 Vaccine. New England Journal of Medicine 2020; 383:2603–15.

4. Baden LR, El Sahly HM, Essink B, et al. Efficacy and Safety of the mRNA-1273 SARS-CoV-2 Vaccine. New England Journal of Medicine 2020; 384:403–16.

5. Sadoff J, Gray G, Vandebosch A, et al. Safety and Efficacy of Single-Dose Ad26.COV2.S Vaccine against Covid-19. N Engl J Med 2021; 384:2187–201.

6. Hungerford D, Cunliffe NA. Real world effectiveness of covid-19 vaccines. BMJ 2021; 374:n2034.

7. Jones JM, Stone M, Sulaeman H, et al. Estimated US Infection-and Vaccine-Induced SARS-CoV-2 Seroprevalence Based on Blood Donations, July 2020-May 2021. JAMA 2021; 326:1400–9.

8. Team CC-VBCI. COVID-19 Vaccine Breakthrough Infections Reported to CDC - United States, January 1-April 30, 2021. MMWR Morb Mortal Wkly Rep 2021; 70:792–3.

9. Jefferson T, Spencer EA, Brassey J, et al. Transmission of Severe Acute Respiratory Syndrome Coronavirus-2 (SARS-CoV-2) from pre and asymptomatic infected individuals: a systematic review. Clin Microbiol Infect 2022; 28:178–89.

10. Tartof SY, Slezak JM, Fischer H, et al. Effectiveness of mRNA BNT162b2 COVID-19 vaccine up to 6 months in a large integrated health system in the USA: a retrospective cohort study. Lancet 2021; 398:1407–16.

11. Butt AA, Omer SB, Yan P, Shaikh OS, Mayr FB. SARS-CoV-2 Vaccine Effectiveness in a High-Risk National Population in a Real-World Setting. Ann Intern Med 2021; 174:1404–8.

12. Andrejko KL, Pry J, Myers JF, et al. Prevention of COVID-19 by mRNA-based vaccines within the general population of California. Clin Infect Dis 2021.

13. Self WH, Tenforde MW, Rhoads JP, et al. Comparative Effectiveness of Moderna, Pfizer-BioNTech, and Janssen (Johnson & Johnson) Vaccines in Preventing COVID-19 Hospitalizations Among Adults Without Immunocompromising Conditions - United States, March-August 2021. MMWR Morb Mortal Wkly Rep 2021; 70:1337–43.

14. Rosenberg ES, Holtgrave DR, Dorabawila V, et al. New COVID-19 Cases and Hospitalizations Among Adults, by Vaccination Status - New York, May 3-July 25, 2021. MMWR Morb Mortal Wkly Rep 2021; 70:1150–5.

15. Tseng HF, Ackerson BK, Luo Y, et al. Effectiveness of mRNA-1273 against SARS-CoV-2 omicron and delta variants. medRxiv 2022.

